# Primary health facility readiness to care for infants under six months at risk of poor growth and development: A HHFA-based survey

**DOI:** 10.1101/2024.04.24.24306298

**Authors:** Tabitha D. van Immerzeel, Abou Ba, Maty Diagne, Indou Deme-Ly, Amanda E. Murungi, Rebecca Penzias, Daouda Seck, Abdallah Diallo, Carlos S. Grijalva-Eternod, Marko Kerac, Louise T. Day

**Affiliations:** Cheikh Anta Diop University, Dakar, Senegal; Department of Infectious Disease Epidemiology and International Health, London School of Hygiene & Tropical Medicine, London, UK; Social Paediatric Institute, Dakar, Senegal; National Paediatric Hospital Albert Royer, Dakar, Senegal; Mulago National Referral Hospital, Kampala, Uganda; Department of Population Health, London School of Hygiene & Tropical Medicine, London, UK

## Abstract

Malnutrition in infants under six months of age (u6m) is poorly identified and managed in many countries, increasing these children’s risk of poor growth and development and preventable mortality and morbidity. New 2023 WHO malnutrition guidelines recommend assessment, classification and treatment at primary health care level. This study aimed to assess primary health facility availability and readiness for WHO-recommended nutritional care in infants u6m.

We adapted the WHO Harmonized Health Facility Assessment (HHFA) with additional items for <u>M</u>anagement of small and nutritionally <u>A</u>t-risk Infants u6m and their <u>M</u>others (MAMI): the HHFA-MAMI tool. Methods included survey of health care providers, direct observation and data extraction from routine registers to calculate mean percentages of a set of items for different readiness areas. We assessed 15 primary health facilities in Senegal, focusing on the five contact points for infants u6m: delivery and postnatal care, immunization programmes, sick child clinics and community health care.

The HHFA-MAMI mean scores (n=15) were: general service availability 51%, general service readiness 69%, management & finance 50%, clinical quality of care 47%. MAMI availability scored 48% and readiness 35%, MAMI infrastructure scored 72%, health workforce 61%, assess MAMI 54%, classify MAMI 15% and treat MAMI 38%, equipment 33% and guidelines & training 22%. Service utilization was highest in postnatal care and immunization contact points, MAMI availability & readiness was highest in delivery and postnatal care.

We conclude that primary health facilities in Senegal have potential to care for infants u6m at risk of poor growth and development with high general PHC readiness and frequent use of care contact points for infants u6m. MAMI availability and readiness require improvements to provide WHO-recommended care.

## Introduction

Infants under six months of age (u6m) are often left behind in global efforts to reduce child malnutrition [1, 2]. Nutritional risk begins in utero, and continues during the neonatal period and thereafter [3]. More than a quarter of babies worldwide are born low birth weight (LBW) either as a result of being preterm, growth-restricted or both [4]. In a review of 54 low and middle-income countries, between birth and six months of age, an estimated 20.1% classified as underweight, 21.3% wasted and 17.6% stunted [5]. When at risk at u6m, these infants will continue to be at risk of both wasting and stunting later in life [6] and have a higher risk of mortality [7]. Longer-term consequences include impaired development [8],[9] and increased risk of developing cardiometabolic non-communicable diseases in adult life [10].

In 2023, the World Health Organization (WHO) published *Guidelines on the prevention and management of wasting and nutritional oedema (acute malnutrition) in infants and children under 5 years,* which propose a transformative shift in approach for infants u6m, from predominantly hospital-based care to outpatient, primary health care (PHC) [11]. The rationale is that improved PHC will reach more infants u6m and prevent adverse outcomes [12]. Defined by WHO as “infants less than six months of age at risk of poor growth and development”, the emphasis of the guidelines is on early detection and timely support for the infant and mother dyad. The case definition for programme enrolment includes infants born small or preterm as well as infants u6m with poor anthropometry as a single or sequential measure (Textbox 1). Treatment, individually tailored for the dyad, includes intensive feeding support (breastfeeding or/and supplementary feeding for lactational failure) and detection and management of physical and mental health conditions for the mother and infant.

### Textbox 1.

#### Summary of WHO 2023 guidelines for infants under six months at risk of poor growth and development

**Summary WHO 2023 guidelines for infants under six months at risk of poor growth and development**

##### Assess

– Screening at care contact points (e.g. delivery care, postnatal care, immunization programs, sick child clinics, community health care);
– Assessment of anthropometry, feeding, infant clinical and maternal nutrition and mental health;
– Mothers and infants are considered an interdependent dyad. Classify
– Infants u6m with poor anthropometry (weight for length z-score <-2SD/ weight for age z-score) <-2SD/ mid upper arm circumference < 110 from 6 weeks) based on a single measure; Infants u6m with poor growth based on sequential measures;
– Infants born preterm/ low birth weight/ small for gestational age;
– Infants with known risk factors for poor growth and development (e.g. feeding problems, congenital issues).

##### Treat

– Referral to inpatient care for any ‘danger signs’ (as per IMCI e.g. fast breathing, lethargic, convulsions, refuses to drink);
– Outpatient treatment focused support for any underlying problem e.g. clinical treatment;
– Breastfeeding support, supplementary feeding (after comprehensive assessment), maternal mental health support;
– Follow-up until six months of age.

A care package consistent with the new WHO guidelines [13] had been previously developed by the MAMI (Management of small and nutritionally At-risk Infants u6m and their Mothers) Global Network of researchers, programmers and policy makers [14]. This MAMI Care Pathway uses similar case definitions and care principles as the WHO guidelines, and has been used to frame and inform our study [15].

Now that countries are starting to implement the new WHO guidelines, health facility assessment is an important first step in adapting to local contexts. WHO recently launched the Harmonized Health Facility Assessment (HHFA) [16], a novel comprehensive tool advancing previous tools. The HHFA data collection uses mixed methods: health worker surveys, direct observations and data extraction from routine registers, using closed questions and checklists. Items of service readiness are measured across the following dimensions and areas:

1. General service availability (areas: health infrastructure, health workforce and services available),
2. General service readiness (areas: amenities, basic equipment, infection prevention, diagnostic capacity, essential medicines and commodities),
3. An optional dimension of Service-specific availability & readiness
4. Management & finance (areas: facility governance, finances and accounting, staff support systems, quality and safety systems, information systems), and
5. Clinical quality of care (areas: record review for specific services).

The concept of service readiness assessment has been used to evaluate various programmes, such as emergency obstetric care [17], non-communicable diseases [18] and elderly care [19]. In the HHFA, a few nutrition related items are captured such as infant scales under equipment and service - specific items for small and sick newborns. However, nutritional care for infants u6m at PHC is currently not captured by the HHFA.

In Senegal, as in many countries, PHC for infants u6m is provided at five contact points along the continuum of care: delivery care, postnatal care, immunization programmes, sick child clinics and community health care [20]. Most (82%) births take place in a health facility [21], providing essential newborn care. Most (80%) mothers attend at least one postnatal visit [21]. Full vaccination coverage at 23 months is currently 77% [21]. Integrated Management of Childhood Illnesses (IMCI) is offered at many primary health facilities [22], but healthcare-seeking behaviour remains low at 50% for fever or diarrhoea [21]. Community preventative services, including growth monitoring activities, are offered for children from birth to 59 months [23]. Currently, infants with low birth weight (estimated prevalence 11%) or malnutrition u6m (estimated prevalence 8%) [24], are managed as inpatients at secondary (health centres) or tertiary level (hospital) according to national guidelines [25]. However, qualitative research revealed mothers’ and health providers’ preference for outpatient care for uncomplicated cases, to make care more accessible and closer to the community [26].

This study aimed to assess primary health facility availability and readiness for WHO-recommended nutritional detection and care in infants under six months. The objectives were:

1. To measure general service readiness in primary health facilities in Senegal
2. To measure service-specific MAMI availability and readiness in primary health facilities in Senegal
3. To assess opportunities to implement WHO-recommended nutritional care for infants u6m at five primary health care contact points.

## Methods

We conducted a descriptive, cross-sectional survey in 15 PHC facilities in three districts in Senegal, from 9 June to 17 September 2023, reported in alignment with the STROBE guidelines [27].

### Setting and sites

In Senegal, primary health facilities are staffed by a team of at least one certified state nurse, one midwife and a few nurse assistants. Each health facility is assigned a catchment area target population and provides all basic primary care for this population, including infants u6m at five contact points: delivery with essential newborn care, postnatal care, immunization programme and sick child clinic (IMCI). Community health care is coordinated by these facilities, each for its catchment areas. The health district provides administrative and financial management and supervision of the primary health facilities but development partners coordinate and finance the community health care. The country of Senegal has 77 health districts and 809 primary health facilities, each covering on average 11,500 inhabitants [28].

### Survey tool development

We adapted the original HHFA [16] (Version 2) for PHC level in Senegal using the national health care policy plan [28], expert review including co-authors who practice clinically in Senegal (TvI, AB and DS), and feedback from pilot testing. Adaptation kept fidelity to the dimensions (general service availability, general service readiness, management & finance, and clinical quality of care) as well as their areas (Figure 1). Sub-areas and items under the sub-areas were selected and adapted applying several iterative steps:

**Figure 1.**
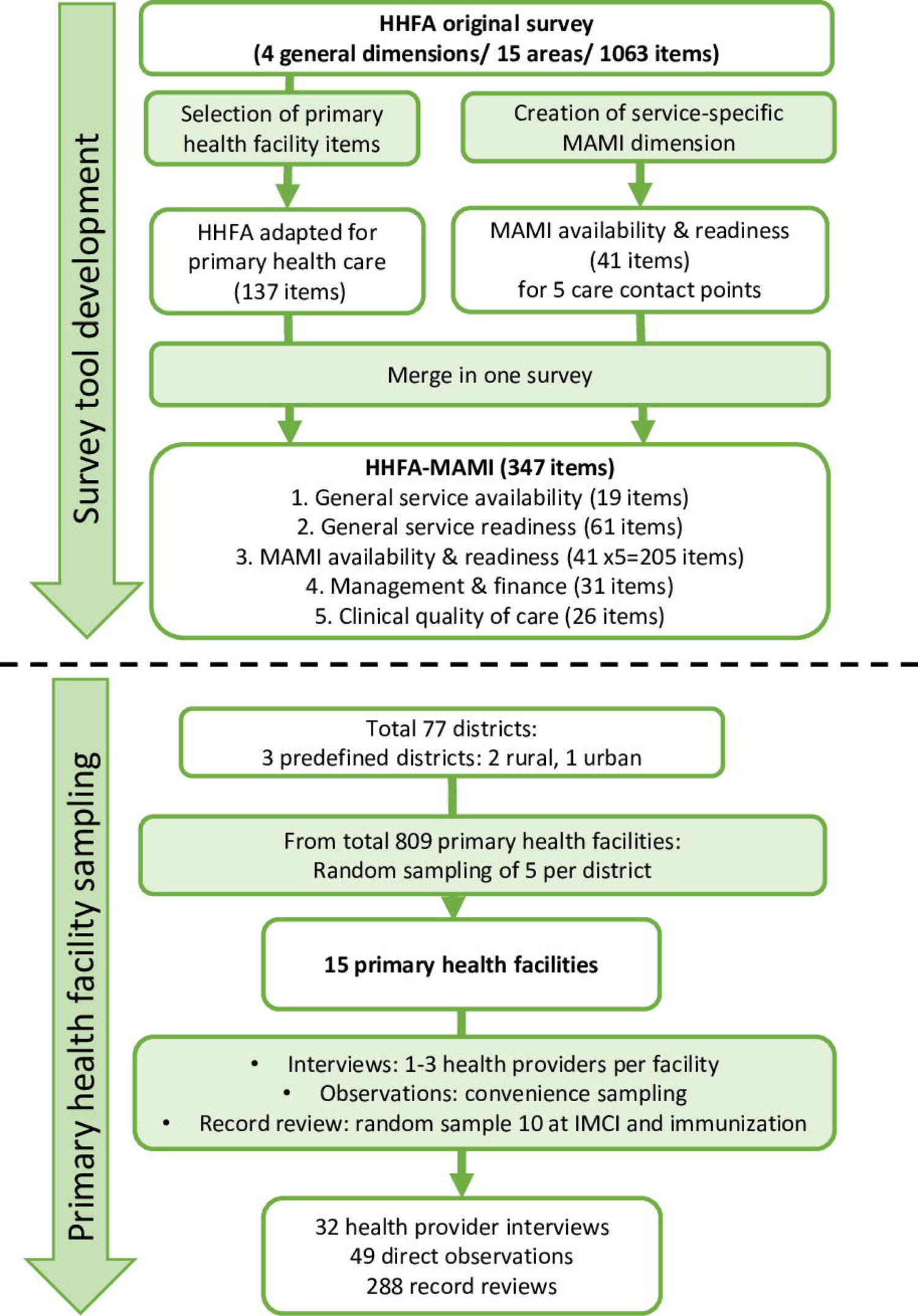
Flow diagram of HHFA-MAMI tool development and health facility survey sampling

– Selecting sub-areas relevant for PHC level (e.g. sub-area “beds” and “oxygen services” were not selected)
– Removing items that are not relevant for primary care in Senegal (e.g. “existence of a quality committee”, which is not implemented on this level).
– Reformulating certain items to be more appropriate to the local context (e.g. changing “governing board” to “health committee”)
– Reorganising the order of items for efficiency (e.g. all observations of the infrastructure/ equipment at the end of the survey, all questions for the midwife grouped)
– Simplifying certain items, such as capturing equipment as present only when functional.
– For the record review sub-area, we selected immunization and IMCI, because of practicality.
– Adding one sub-area: to complete the clinical quality of care dimension, the sub-area “direct observations of care” was added. We used the observation checklist from SARA (Service Availability and Readiness Assessment)[29].

For the health facility density and nurse/ midwife density we used the WHO global standards [30].

Following the format of the original HHFA, we collected three primary data:

– Health care provider survey closed questions, (yes/ no, or checklist), used for items concerning service delivery and organization of care.
– Direct observations, using a checklist for observed items (used for building, equipment, commodities and hygiene).
– Data from registers and reports, used for record review, number or patient visits and target population.
The final primary health facility assessment tool consisted of a selection of 137 of the 2571 original items across the four dimensions (S1 Appendix 1 for a full list of items).
We created a service-specific dimension “MAMI availability & readiness”, using similar areas as other service-specific dimensions that exist in the HHFA: health infrastructure, health workforce, service availability, equipment and guidelines & training. We (TvI, AB and LD all clinicians and public health practitioners) mapped items from the evidence-based MAMI Care Pathway [14], developed in 2021 which aligns with the 2023 WHO guidelines. The MAMI Care Pathway uses the WHO IMCI structure of “assess, classify and treat”, which we adopted as sub-headings under MAMI availability. The final “MAMI availability & readiness” dimension had 41 items, repeated for five care contact points, making 205 items in total (Figure 1, Annex 1).

The draft HHFA-MAMI tool was translated into French and pilot tested by TvI and a research assistant in a non-participating health facility and further revised by an iterative process in two rounds.

### Sampling

We used a multi-step sampling process (Figure 1). First, we purposively selected three districts among the 77 in Senegal-where collaborators had existing partnerships: two rural (Dahra, Kaffrine) and one urban (Pikine) district. The sample size was determined by resources available for the study, as has been described in earlier service readiness studies with sample sizes of between 12 and 30 health facilities [31],[32]. We selected fifteen PHC facilities, five from each district using random number allocation from the sample frame list from the chief medical officer (18 facilities in Dahra, 24 in Kaffrine and 12 in Pikine). This was a descriptive pilot study, not testing hypotheses, therefore we did not calculate formal a-priori sample size.

### Data collection

Two researchers (TvI - a Dutch medical doctor living and working in Senegal and a Senegalese state nurse) conducted the HHFA-MAMI survey over two days at each health facility. Participants were recruited from the selected facilities from 9 June to 17 September 2023. On the first day, a face-to-face general questionnaire was administered to the focal point of the facility– the head nurse, who chose the convenient time and place. The head nurse then provided access to reports and registers needed for data extraction, appointed staff members with whom to conduct the rest of the face-to-face survey and organised the observations. The following day was used for observations of building, equipment and consultations and the remaining survey questions. We recruited mother/ infant dyads for observing a consultation at convenience (all infants u6m consulted at any of the contact points during the second survey day). Register data were accessed during the same time period (from 9 June to 17 September 2023), record reviews were selected randomly (ten from each contact point, starting at a different month at each facility).

### Data management and analysis

HHFA-MAMI data were collected using a password-protected tablet onto a customized digital ODK form [33]. The tablet synchronised with an encrypted central database, manually checked by the data collection team, exported [34] and saved on the LSHTM-secured cloud storage, on a password-protected device. We calculated the mean percentage of health facilities (n=15) that scored positively for each item (Figure 1), and the mean scores of a group of items (sub-area), with equal weighting. An area as well as dimension total mean score was calculated, again with equal weighting. These scores were used to compare between the five contact points.

### Ethics

Ethical approval was obtained from the National Ethics Committee in Senegal (Ref: SEN 19/78) and LSHTM (Ref: 28311). Written informed consent was obtained from all health providers with provision of an information sheet prior to conducting the surveys. Written informed consent with provision of an information sheet, was obtained from the mothers, or a trusted witness (family member/ neighbour) at the mother’s request, before observing a consultation with their infant. Although patient identifiable data were accessible to the researchers who extracted data, all register data were collected with no identifiers.

## Results

The catchment area population for the included primary health facilities was (median, interquartile range (IQR)) 14,557 (9079-26,098) people of which 320 (213-415) infants u6m (Table 1).

**Table 1.** Characteristics of the population being served and seeking care at the five care contact points of the selected PHC facilities.

| Population characteristics (2022) for selected primary health facilities (n=15) | (median, IQR) |
| --- | --- |
| Target whole population | 14,557 (9079-26,098) |
| Expected births | 596 (373-963) |
| Target population u6m | 320 (213-415) |
| Service utilisation (number of visits in 2022) |  |
| Curative care visits all ages (n=15) | 4202 (2990-9190) |
| Delivery care (n=15) | 295 (151-410) |
| Postnatal care (n=15) | 578 (315-1319) |
| Immunization program (n=15) | 719 (389-1541) |
| Sick child clinic (n=15) | 216 (133-419) |
| Community Health (n=8) | 248 (102-482) |

In each of the 15 primary health facilities, we conducted between one and four interviews (n=32), zero to five direct care observations (n=49) and 19-20 record reviews (n=288).

## General service readiness in primary health facilities (objective 1)

The HHFA-MAMI general scores (n=15) were: general service availability 51%, general service readiness 69%, management & finance 50% and clinical quality of care 47%.

Within general service availability, infrastructure (50%) and health workforce (21%) scored lowest with only four facilities meeting the primary health facility density indicator and one meeting the nurse/ midwife density indicator (Table 2).

**Table 2.**
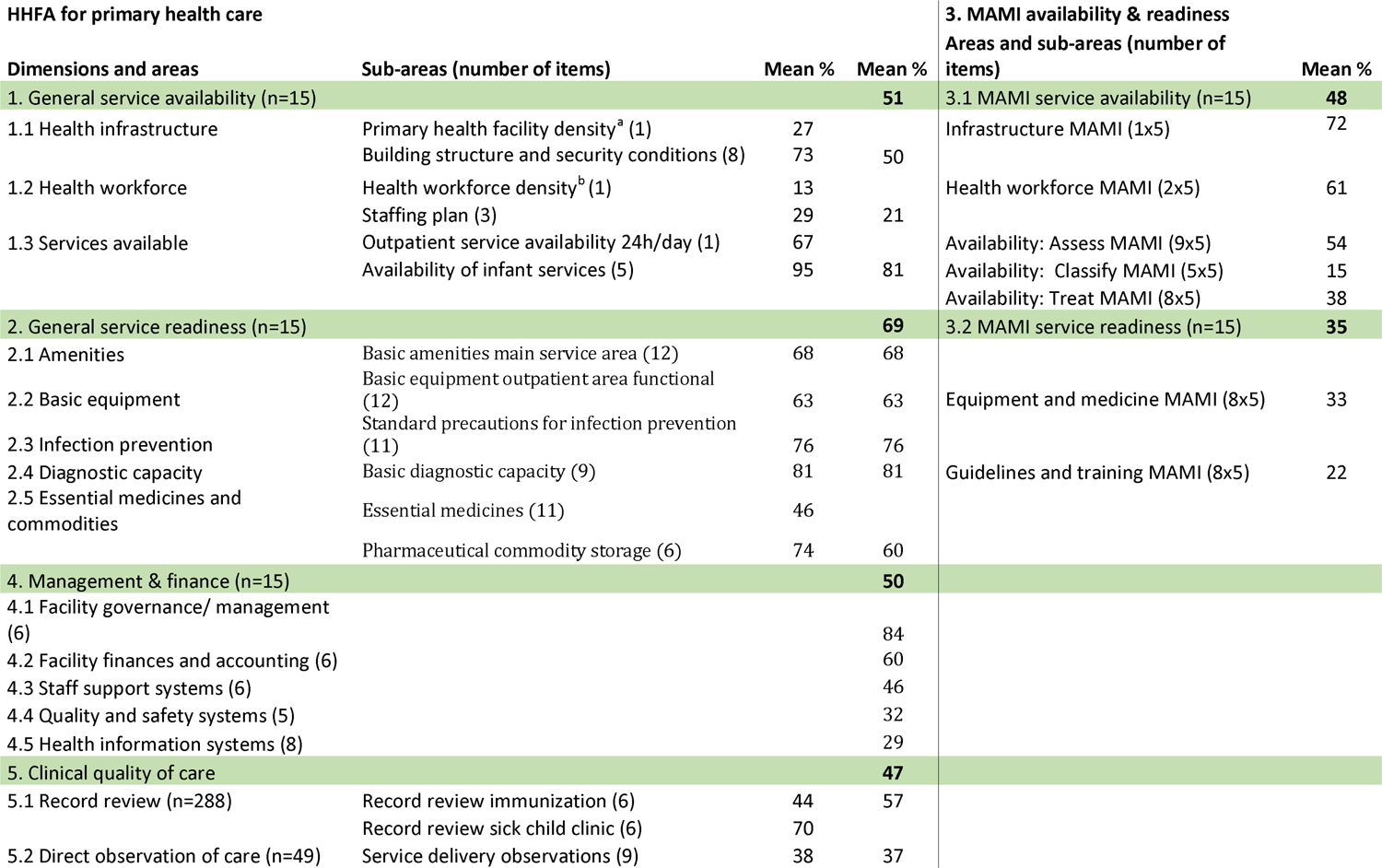
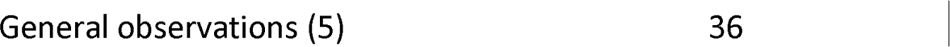
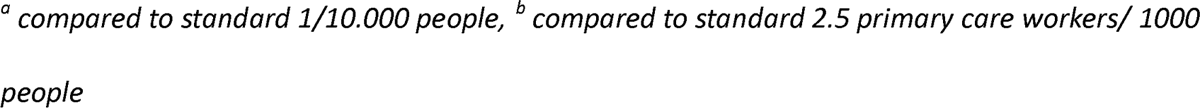
HHFA-MAMI results: primary health care dimensions and MAMI availability & readiness dimension.

Within general service readiness, amenities scored 68%, basic equipment 63%, infection prevention 76%, diagnostics capacity 81% and essential medicines and commodities 60%. Within management & finance, governance (84%) and facility finances (60%) scored higher than staff support systems (46%), quality and safety systems (32%) and health information systems (29%). Clinical quality of care scored 57% on record review and 37% on direct observations of care. (Table 2)

### Service-specific MAMI availability & readiness (objective 2)

Overall, the MAMI availability & readiness dimension scored 48% on availability and 35% on readiness. Infrastructure 72% and health workforce 61% scored higher than availability assess MAMI 54%, classify MAMI 15% and treat MAMI 38%. Equipment scored 33% and guidelines & training 22% (Table 2).

Service utilisation (median patient visits in 2022, IQR) was high in immunization programmes (719, 389-1541) and postnatal care (578, 315-1319), compared to a median of 4202 (2990, 9190) patient curative visits in 2022 for all ages (Table 1). Scores per sub-area with mean service utilisation for each care contact point are shows in Figure 2.

**Figure 2.**
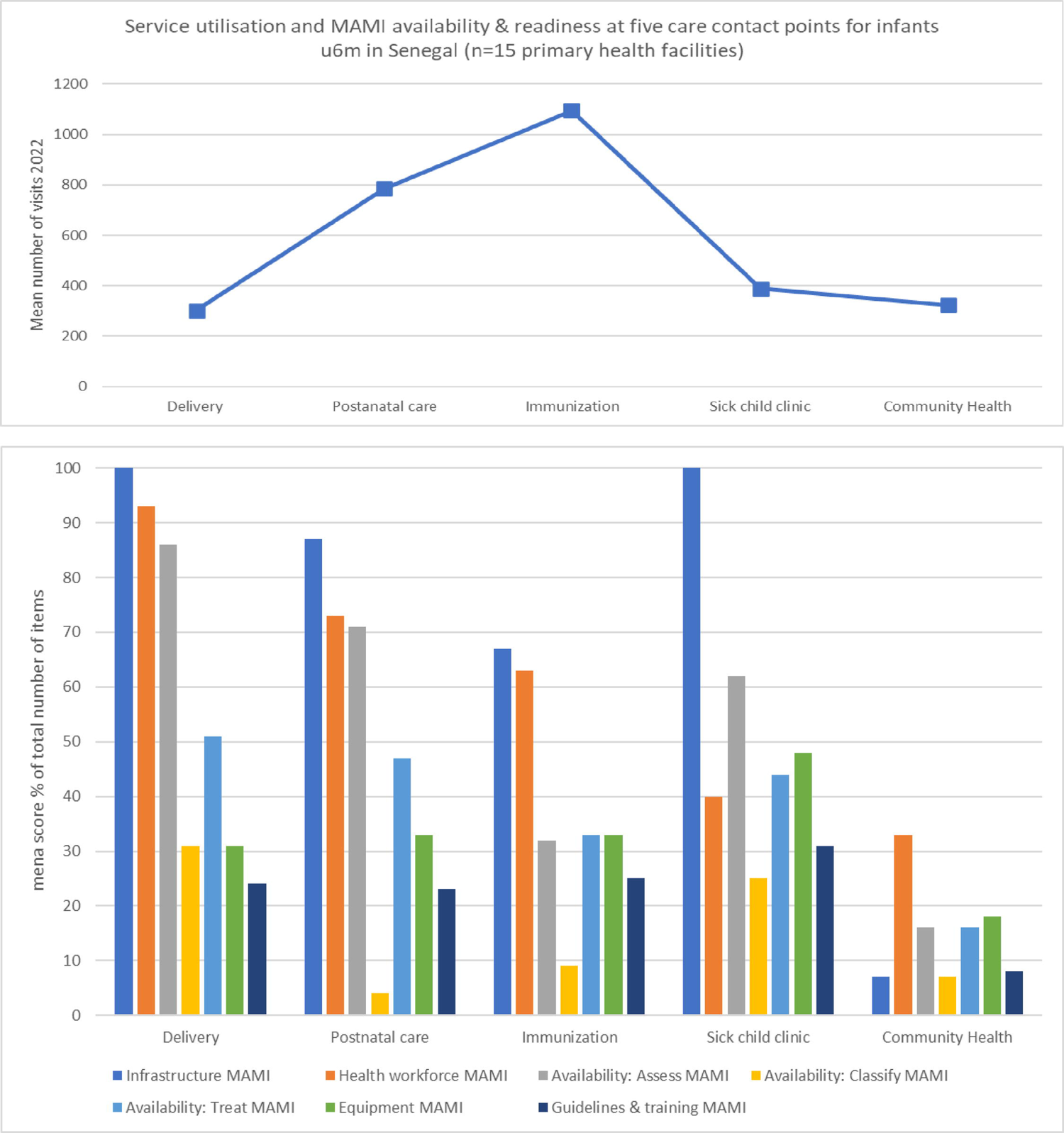
HHFA-MAMI assessment of mean service utilisation and availability & readiness for five care contact points in three districts in Senegal (n=15 PHC facilities)

### Nutritional care for infants u6m at five primary health care contact points (objective 3)

Comparisons of scores between the five contact points are visualised in a heatmap in Figure 3.

**Figure 3.**
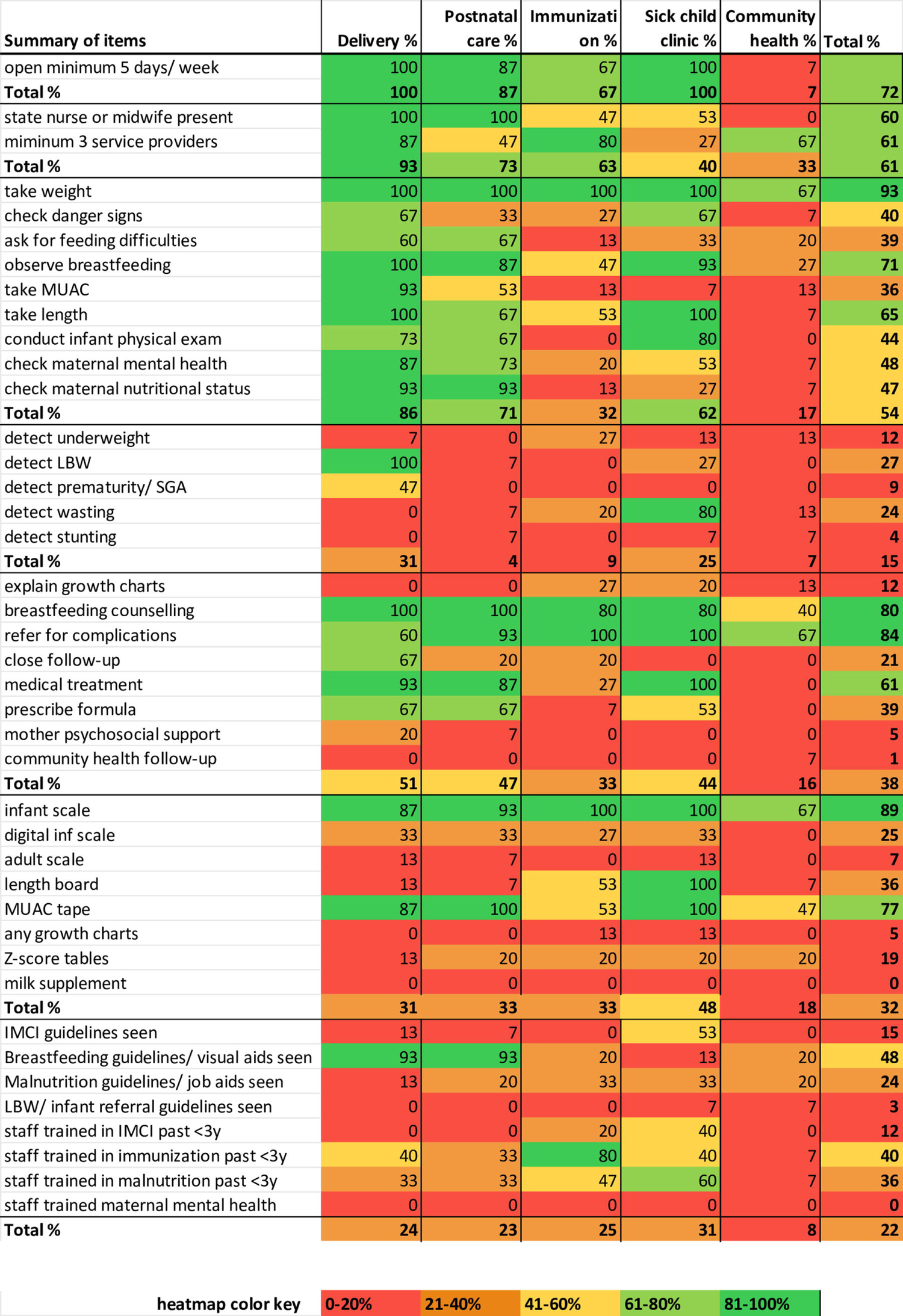
Heat map MAMI availability & readiness items for five care contact points for infants u6m (n=15 PHC facilities)

Infrastructure for MAMI scored high (72%): Delivery, postnatal care and sick child clinics were mostly open seven days a week, while immunization programmes were often provided weekly. Community health activities were provided monthly and were only offered in the two rural districts.

Health workforce, defined by the number of health providers available as well as their level of training, scored 61%. Delivery care, postnatal care and sick child clinics were provided by midwives or state nurses, while immunization service and community health were provided by lower trained health providers. Numbers of health providers were lowest in sick child clinics (27% with three health providers or more), while community health had most service providers (13 on average).

MAMI “Assess”, including measurements such as weight, length or mid-upper-arm-circumference (MUAC), scored higher (54%) than “Classify” (15%) and “Treat” (38%). Health care providers reported that weight was taken at all clinical contact points, while only at delivery care a detailed anthropometric assessment was done, including weight and length (100% of facilities) and MUAC (93%). Midwives routinely observed breastfeeding after birth (100%), but not all of them reported to ask for feeding difficulties (60%). At most care contact points, either breastfeeding observation or asking for feeding difficulties was reported. Most (87%) midwives also reported conducting assessment of maternal nutrition status and maternal mental health, although the survey did not provide detailed information on how this was done. Infant physical examination was reported to be done by midwives (73% of the facilities at delivery, 67% at postnatal care) and providers at sick child clinics (80%), not at immunization programmes or in community health care.

MAMI “Classify” scored low overall (15%), meaning interpreting nutritional deficits with the usage of growth charts and classifying into for example normal/ moderate/ severe malnutrition. Midwives reported diagnosing low birth weight (100%) and sometimes prematurity (47%), while at sick child clinics infants were classified as wasted/non-wasted (80%) using weight for length, rarely MUAC (7%). At immunization programmes, weight was sometimes plotted for age (27%) or for length (20%). At postnatal care and community health, classification was rarely done.

MAMI “Treat” reported referral of at risk infants in 84% of all five contact points at the 15 facilities, either to a different facility contact point (e.g. sick child clinic) or externally to the nearest district hospital or a tertiary centre (up to 300 kilometres away). Breastfeeding counselling was reported to happen at delivery and postnatal care (100%), immunization programmes (80%) and sick child clinics (80%). At over two third (67%) of the delivery and postnatal care contacts points and over half (53%) of the sick child clinics, providers said to regularly prescribe infant formula, for example in case of low birth weight or when the mother self-report insufficient milk.

Explanation of growth charts to the mother was reported to happen at 27% of immunization programmes and 20% of sick child clinics. Medical treatment for infants with clinical conditions was provided at birth (93%), during postnatal care (87%) and during sick child clinics (100%). Community follow-up was mostly absent, with one exception where a midwife reported referral to community health workers for postnatal home visits.

Overall, MAMI equipment readiness at care contact points was 33%. In 93% of the facilities, an adult scale was seen at the general outpatient area, only in 7% of the five contact points for infants u6m. All facilities had at least one functioning infant scale. Most (13) facilities had a functional analogue scale, five a digital, five a SECA-876 mother/ baby scale and at 12 a Salter scale. Health providers at most (12) facilities stated that their equipment is usually calibrated.

A length board was observed in all 15 health facilities. This standardised UNICEF device was seen in sick child clinics (100%) and immunization programs (53%). A standardised coloured MUAC tape was seen in 100% of sick child clinics and 53% of immunization programmes. At delivery care and postnatal care an unrecommended simple measuring tape was seen, used for taking both length and MUAC. Growth charts for weight and length were observed in 5% of the total contact points, although these were present in parent-held immunization cards which were not observed for this study. Laminated weight for height Z-score tables were observed, at consultation desks in postnatal care or sick child clinics (20%). In community health care in the two rural districts we saw a customised “look-up table” developed by the national nutrition advisory board (CNDN) to determine expected monthly weight gain (independent of the infants’ age) [35]. Infant formula milk was not observed to be stocked at any of the facilities.

Guidelines and training relevant for MAMI scored low overall (22%). In over half (53%) of the sick child clinics, IMCI-related guidelines were observed, often displayed on walls. Breastfeeding visual aids existed in the mother/ child record booklets, produced by the ministry of health and midwives reported using them widely for breastfeeding counselling after birth (93%) and during postnatal care (93%). Staff training at contact points over the previous three years was most frequently for immunization (40%), malnutrition (36%) and IMCI (12%). Guidelines or criteria for referral of infants with low birth weight or malnutrition were not seen.

## Discussion

Primary health care facilities in Senegal have potential to care for infants u6m at risk of poor growth and development and their mothers. The HHFA-MAMI survey in 15 facilities found MAMI availability of 48% and MAMI readiness of 35%. This is an important indication that the implementation of 2023 WHO recommendations at PHC level is possible. Infrastructure, health workforce and equipment were available, but varied between service contact points. Among the three steps, infants were frequently assessed including for anthropometry, but not often classified as at risk, which is essential for adequate treatment as outpatients and referral for complications. Breastfeeding support was commonly provided, while the frequent prescription of infant formula showed a need for supplementary feeding guidance. Maternal mental health support was an observed gap. We found opportunities at each of the five contact points to implement WHO nutritional recommendations for at risk infants u6m.

### Interpretation

The HHFA has been created to evaluate implementation of existing health policies, however, we found our HHFA-MAMI adaptation useful and potentially informative at the pre-implementation phase of WHO guidelines for infants u6m at risk of poor growth and development in Senegal. Current low scores could rapidly improve once the new WHO recommendations are formally rolled out and the survey is repeated in the future. Repeated readiness analysis has been used for evaluation purposes in more general maternal and child services in Senegal [36]. Similarly, for small and sick newborns, theNEST360 alliance has developed a readiness tool for repeated analysis of hospital care [37].

Among the four general HHFA dimensions, general service readiness is most used in other studies. Our HHFA-MAMI general service readiness of 69% was slightly higher than other studies that assessed PHC readiness: 60.5% in Bangladesh and 61.5% in Mongolia [38],[39]. Our HHFA general service availability showed neither facility density nor health worker density meeting WHO standards in Senegal, which are important indicators in improving primary care coverage [40]. The dimension management & finance was rarely assessed in other studies, while in a study on elderly care readiness, these aspects performed poorest [19]. Our low scores for health information systems (29%) and quality systems (32%) might need attention, implementing new services. The clinical quality of care dimension is essential in the HHFA, because readiness measured by only structural inputs has shown to poorly correlate with quality of care [41].

Previous nutrition service availability and readiness research for a wider age range (under 5 years) used similar items to our HHFA-MAMI tool, e.g. in primary health facilities in Nigeria: “offers child growth monitoring” (91.5%) and “diagnoses child malnutrition” (81.4%) [42]. A multi-country (n=11) nutrition-related readiness analysis at primary care facilities found overall low scores, comparable to our study: 30% “counselled on child growth” and 21%” plotted weight in growth chart” [43]. Stakeholder consultation to developed a readiness indicator list focussed on nutritional quality of care for mothers and children under five years showed similarities to our tool, such as items “ask for feeding”, and “plotting the weight in a growth curve” [44].

Although our data did not allow us to calculate service coverage, the service utilisation gave some indication of how many infants and mothers are seen at the contact points. Combining service utilisation with readiness had earlier been described as a useful way to assess effective coverage [45].

### Actionable findings to implement WHO guidelines at PHC contact points in Senegal

At childbirth services in Senegal, midwives routinely measure birth weight, as in other contexts [46], and in line with the new WHO recommendations for at risk infants u6m, also measure length and MUAC, although some with unrecommended measuring tools.

At postnatal care, breastfeeding support was routinely provided in our study facilities, and extending this practice beyond the postnatal age up to 6m would be crucial for WHO guideline implementation [47]. A standardized breastfeeding assessment tool could help identify those at risk, needing closer breastfeeding support and follow-up [48]. Infant formula was reported to be frequently prescribed in our study although not observed to be stocked in the health facilities and new WHO guidelines could support in supplementary feeding counselling.

Immunization services are expected to monitor children’s growth according to Senegal health policy yet do not always have the anticipated impact for at risk infants [49], due to a lack of proper use of growth charts [50]. There is an urgent need for better tools or lookup tables to simplify the usage of growth charts [51]. Besides weight, other anthropometric measurements are promising in infants u6m such as MUAC, although more evidence is needed to validate cut-off values [52] and to standardize MUAC tapes [53].

Sick child clinics in Senegal apply IMCI although concerns have been expressed regarding fidelity to guidelines [36]. The MAMI Care Pathway uses the IMCI structure “assess, classify, treat”, and aligning WHO guidelines implementation to IMCI would be beneficial. Maternal mental health support is an important element in the new WHO guideline that complements IMCI and could make a major contribution to improving nutritional outcomes for u6m children [54].

Community health care provides opportunities in detecting at risk infants, with larger numbers of health providers than at other care contact points in our study. To benefit from its advantages, task allocation is crucial [55] and there is a need for supervision and clinical mentoring to strengthen this service [56].

### Limitations

Our research is the first effort using the HHFA for u6m infant nutrition assessment, making it novel, though less comparable to similar studies using older tools, which is a limitation. Another limitation is that cut-off points for readiness scores have not yet been defined: what overall score is ‘good enough’ for a particular facility. Future research is needed to validate the HHFA for various settings. However, our HHFA-MAMI tool showed proof of concept in this study and could be tested and validated in wider context.

The HHFA-MAMI captures mainly nutritional items, that do not necessarily cover all the risk factors underlying infant malnutrition [57]. However, a strength of our tool is its parsimonious collection of 137 general and 41 MAMI items from PHC facilities, that require only 1-2 days of data collection by one or two people.

Finally, we acknowledge that our sample size was small to compare scores between districts. However, our data are critical to inform sample size calculations and logistical considerations for future more analytical service-readiness studies.

### Generalisability

A recent stakeholder consultation in 42 countries revealed a perceived need for improved detection and care for at risk infants u6m [58], which will be accelerated by the 2023 WHO malnutrition guidelines. Senegal was mentioned as an exemplar country in stunting reduction, because of its effective implementation of nutrition policy and multisectoral approach [59]. The HHFA-MAMI survey tool we developed and future MAMI implementation lessons learned from Senegal might therefore inspire countries in the region and beyond.

## Conclusion

Primary health facilities in Senegal have potential to care for infants u6m at risk of poor growth and development with high general PHC readiness and frequent use of care contact points for infants u6m (delivery and postnatal care, immunization programmes, sick child clinics and community health care). Our HHFA-MAMI survey found suboptimal MAMI availability and readiness, but many items are in place or could be strengthened, implementing WHO-recommended nutritional care for at risk infants.

## Data Availability

The complete list of items of our HHFA-MAMI survey tool is available in Supplementary file 1. The survey and anonymised results presented in this study are available from the LSHTM data repository on request with the corresponding author (https://datacompass.lshtm.ac.uk/).

https://datacompass.lshtm.ac.uk/cgi/users/home?screen=EPrint::View&eprintid=3819#t

## Acknowledgements

We greatly thank all health care providers we interviewed, who welcomed us in their working place and homes and were willing to share information. We thank the mothers of the infants who allowed us to observe their consultation and whose information we extracted from registers and reports. We thank professor Babacar Faye for facilitating ethics application and professor Saliou Diouf and Johan Velema for their advice on the project. We thank the administrative heads who facilitated the study in each of the three districts in Senegal. A huge thanks to Sheikh Jallow who assisted in data collection. “Electronic data solutions were provided by LSHTM Global Health Analytics (odk.lshtm.ac.uk).”

## Supporting information captions

S1 Appendix. HHFA-MAMI tool complete list of items (English and French)

## Notes

### Competing Interest Statement

The authors have declared no competing interest.

### Funding Statement

The principal investigator received a small grant from Otto Kranendonk Fonds, the Netherlands. The funders had no role in study design, data collection and analysis, decision to publish, or preparation of the manuscript.

### Author Declarations

Ethical approval was obtained from the National Ethics Committee in Senegal (Ref: SEN 19/78) and LSHTM (Ref: 28311).

### Summary of Updates

removed inter quartile range from the results

